# Associations between early in-hospital medications and the development of delirium in patients with stroke

**DOI:** 10.1101/2023.04.27.23288875

**Authors:** Sophia L. Ryan, Xiu Liu, Vanessa McKenna, Manohar Ghanta, Carlos Muniz, Rachel Renwick, M. Brandon Westover, Eyal Y. Kimchi

**Affiliations:** Department of Neurology, Massachusetts General Hospital; Department of Neurology, Mount Sinai Medical Center; Lawrence Center for Quality and Safety, Massachusetts General Hospital; Department of Neurology, SUNY Upstate Medical University; Department of Neurology, Beth Israel Deaconess Medical Center; Department of Neurology, Northwestern University

**Keywords:** stroke, delirium, medications, polypharmacy

## Abstract

**Objectives:** Patients hospitalized with stroke develop delirium at higher rates than general hospitalized patients. While several medications are associated with existing delirium, it is unknown whether early medication exposures are associated with subsequent delirium in patients with stroke. Additionally, it is unknown whether delirium identification is associated with changes in the prescription of these medications.

**Materials and Methods:** We conducted a retrospective cohort study of patients admitted to a comprehensive stroke center, who were assessed for delirium by trained nursing staff during clinical care. We analyzed exposures to multiple medication classes in the first 48 hours of admission, and compared them between patients who developed delirium >48 hours after admission and those who never developed delirium. Statistical analysis was performed using univariate testing and multivariable logistic regression.

**Results:** 1,710 unique patients were included in the cohort, of whom 471 (27.5%) developed delirium >48 hours after admission. Delirium was associated with prior exposure to antipsychotics, sedatives, opiates, and antimicrobials, even after accounting for several clinical covariates. Usage of these medications decreased in the 48 hours following delirium identification, except for atypical antipsychotics, whose use increased. Other medication classes such as steroids, benzodiazepines, and sleep aids were not initially associated with subsequent delirium, but prescription patterns still changed after delirium identification.

**Conclusions:** Early exposure to multiple medication classes is associated with the subsequent development of delirium in patients with stroke. Additionally, prescription patterns changed following delirium identification, suggesting that these medication classes may be modifiable targets for future delirium prevention strategies.

**140-character message:** Early exposure to specific medication classes is associated with delirium in patients with stroke & may be targets for future prevention.

@SophiaLRyan, @ekimchi, @MGHNeurology, @MSHSNeurology, @NMNeurology

## INTRODUCTION

Delirium is a disorder of attention and awareness that is common among patients with stroke and is associated with a longer length of stay, increased risk of readmission, and death.^1–3^ Patients with stroke are particularly vulnerable to delirium compared to the general hospitalized population (13-48% occurrence in patients with stroke, compared to 10-25% occurrence among the general hospital population).^1^ Given that delirium is often incident, arising after hospitalization,^4^ it is important to identify factors that may affect or anticipate its development.^5^ Delirium can be prevented through nonpharmacologic interventions in general medical^6^ and general neurology^7^ inpatients, however, it is less clear whether such interventions can prevent or treat delirium in patients with stroke.^8–10^ The prevention and management of delirium remain an important challenge in the care of hospitalized patients with stroke.

Delirium has many potential contributing factors, both non-modifiable factors, such as age and cognitive impairment,^11–16^ and potentially modifiable factors, including medications.^17–19^ In addition to polypharmacy, certain classes of medications are thought to be deliriogenic including sedatives,^20^ antimicrobials,^21, 22^ opiates,^23^ antiepileptics,^24^ medications for the treatment of Parkinson’s disease,^25–27^ steroids^28, 29^, benzodiazepines,^30^ anticholinergics,^31, 32^ antiemetics,^33^ mood stabilizers^34^ and muscle relaxants.^35^ Avoidance of potentially deliriogenic mediations, where clinically appropriate, may represent an opportunity for delirium prevention.

While associations between in-hospital medications and delirium have been examined in older patients,^36, 37^ ICU patients,^38, 39^ and post-operative patients,^30^ it is not currently known how medications administered early in a hospitalization impact the subsequent development of delirium in patients with stroke. Additionally, there is no literature describing to what extent the administration of such medications may be clinically modifiable once delirium has occurred. We therefore conducted a retrospective cohort study to determine which medications administered in the first 48 hours of hospitalization predicted the subsequent development of delirium in patients with stroke. Additionally, we sought to characterize how prescribing patterns changed after delirium was identified.

## METHODS

### Data availability

Anonymized data not published within this article will be made available by request from qualified investigators.

### Patient population

We performed a retrospective cohort study of all patients admitted to our comprehensive stroke center with acute neurovascular disease based on ICD10 coding, including ischemic or hemorrhagic stroke, between January 2017 and April 2019. Given that we were interested in the effect of medications administered within the first 48 hours of hospitalization on the subsequent development of delirium, only those with a hospital stay of greater than 48 hours and a recorded delirium screen were included. We excluded patients who screened positive for delirium in the first 48 hours and those who did not have any delirium screen recorded. Only one admission was included per patient. The sample size was determined by the number of admissions during this time as a sample of convenience. Race, ethnicity, marital status, English speaking and insurance payor were all obtained from an internal quality improvement database.

### Clinical data

Delirium assessments were extracted from the electronic medical record, in addition to patient demographics, baseline characteristics, medication administration, and clinical outcomes. When available, the NIH stroke scale (NIHSS) was also extracted. We used admission nursing screens to determine rates of visual and hearing impairment, denture use, dependence in activities of daily living, and memory impairment. Potentially hazardous alcohol use was determined by either an Audit C score on admission suggestive of alcohol overuse^40^ or use of a Clinical Institute Withdrawal Assessment (CIWA) scale.^41^ We also collected internal administrative data identify neurovascular intervention. Relative costs were determined from a hospital administrative database in relative units, as per hospital administrative policy.

### Delirium assessment

The primary outcome of this study was the development of delirium 48 hours or more after admission. Delirium was assessed by trained neurology nurses once per shift (twice daily) as part of routine clinical care, using the CAM-ICU in the Intensive Care Unit^42, 43^ and a Confusion Assessment Method (CAM) framework on the wards^44^. CAM-based algorithms for delirium identification have been validated in various patient populations, including the general hospital population^44, 45^ and patients with acute stroke^2, 43, 46^.

Nursing staff were trained to perform assessments as part of a departmental quality initiative through a combination of in-person didactics, in-service sessions, online training, and targeted feedback including ongoing discussions from nurse supervisors. Nurses were specifically trained regarding CAM administration in the neurosciences population, including in the setting of aphasia and dementia. Staff could discuss complex or uncertain cases during daily interdisciplinary group rounds, where formalized interdisciplinary checklists prompted the identification of each patient’s delirium status.

Patients were stratified by their delirium status into those who never had a positive delirium assessment and those who first screened positive >48 hours after admission. We excluded patients who developed delirium within the first 48 hours of hospitalization, in order to examine the importance of potentially delirious medications prior to delirium onset.

### Medication data

The main exposures in this study were selected medication classes. Based on the literature and *a priori* clinical knowledge, we selected the following potentially deliriogenic medication classes for study: sedatives,^20^ antimicrobials,^21, 22^ opiates,^23^ antiepileptics,^24^ medications for the treatment of Parkinson’s disease,^25–27^ steroids^28, 29^, benzodiazepines,^30^ anticholinergics,^31, 32^ antiemetics,^33^ mood stabilizers^34^ and muscle relaxants.^35^ We also looked at sleep aids, as these are often used to manage the disrupted sleep-wake cycles of delirious patients,^47, 48^ stimulants, which may cause hyperactive delirium and are sometimes used off-label for the treatment of hypoactive delirium,^49–51^ and cholinesterase inhibitors and memantine,^52, 53^ which have been trialed in the prevention and treatment of delirium although data has been mixed. Finally, we also examined antipsychotic medications, which are frequently used off-label in the management of delirium but may also be potentially deliriogenic.^54, 55^

Anticholinergic medications were defined according to the American Geriatric Society’s 2019 Beers criteria.^19^ We restricted each medication to appear in only a single therapeutic class based on clinical knowledge of common usage (e.g. quetiapine as an atypical antipsychotic, rather than also an anticholinergic). Medications in each class are listed in Supplemental Table S1. Lastly, when medications were administered via routes with minimal systemic absorption they were removed (i.e. topical, inhaled, eye or ear drops, etc).

### Statistical analysis

Descriptive analyses were used to summarize the characteristics of patients who did and did not develop delirium in terms of (1) demographic and baseline characteristics; (2) clinical characteristics related to their hospitalization; (3) clinical outcomes. Medians and interquartile ranges were presented for continuous variables, and percentages were calculated for categorical variables.

We compared medication class exposures during the first 48 hours of admission between patients who did and did not developed delirium. Due to variability in the medications used, including doses and routes of administration, as well as a lack of consensus on dose conversion for several classes, we evaluated medication exposures rather than doses. Chi-square or Fisher’s exact tests were used for categorical variables and the Wilcoxon rank sum test was used for continuous variables.

Based on the univariate analysis and *a priori* knowledge, we built a multivariable logistic regression model controlling for potential confounders and significant predictors of delirium (including hemorrhagic stroke, ventilator status during the first 48 hours, age, and sex) along with exposures to univariate statistically significant medications. Ventilator status instead of ICU status was utilized to control for its strong association with sedatives. Risk ratios and 95% confidence interval were presented in the multivariable regression analysis

Finally, among those patients who developed delirium, we compared the change in proportions of patients receiving potentially deliriogenic medications in the 48 hours before and after delirium identification, using McNemar’s test. A p-value < 0.05 was considered statistically significant in all analyses. Descriptive statistics were performed using MATLAB (R2018b, MathWorks), and inferential statistics were performed using SAS software, version 9.4 (SAS Institute).

### Approval

This study was approved by the Institutional Review Board at Massachusetts General Hospital (Boston).

## RESULTS

### Patient demographics and baseline characteristics

12,228 admissions to the clinical neuroscience wards occurred during the study period, of whom 1,710 unique patients with acute neurovascular diagnoses were included for analysis (Figure 1). Of these, 1,020 patients presented with primarily ischemic stroke and 649 patients with primarily intracranial hemorrhage. The remaining 41 patients had other symptomatic neurovascular compromise (for example, MRI negative transient ischemic attack). Delirium developed 48 hours or more after admission in 27.5% of patients (471/1710; Table 1). Patients who developed delirium 48 hours or more into their hospitalization, as compared to those who did not develop delirium, tended to be older, have Medicare or Medicaid insurance, have pre-existing hearing and memory impairment, and required assistance with more activities of daily living (ADLs). Sex, ethnicity, race, and English fluency were similar between patients with and without delirium. Patients who were not assessed for delirium during their hospitalization tended to be younger and have shorter lengths of stay (Supplementary Table S2).

**Figure 1.**
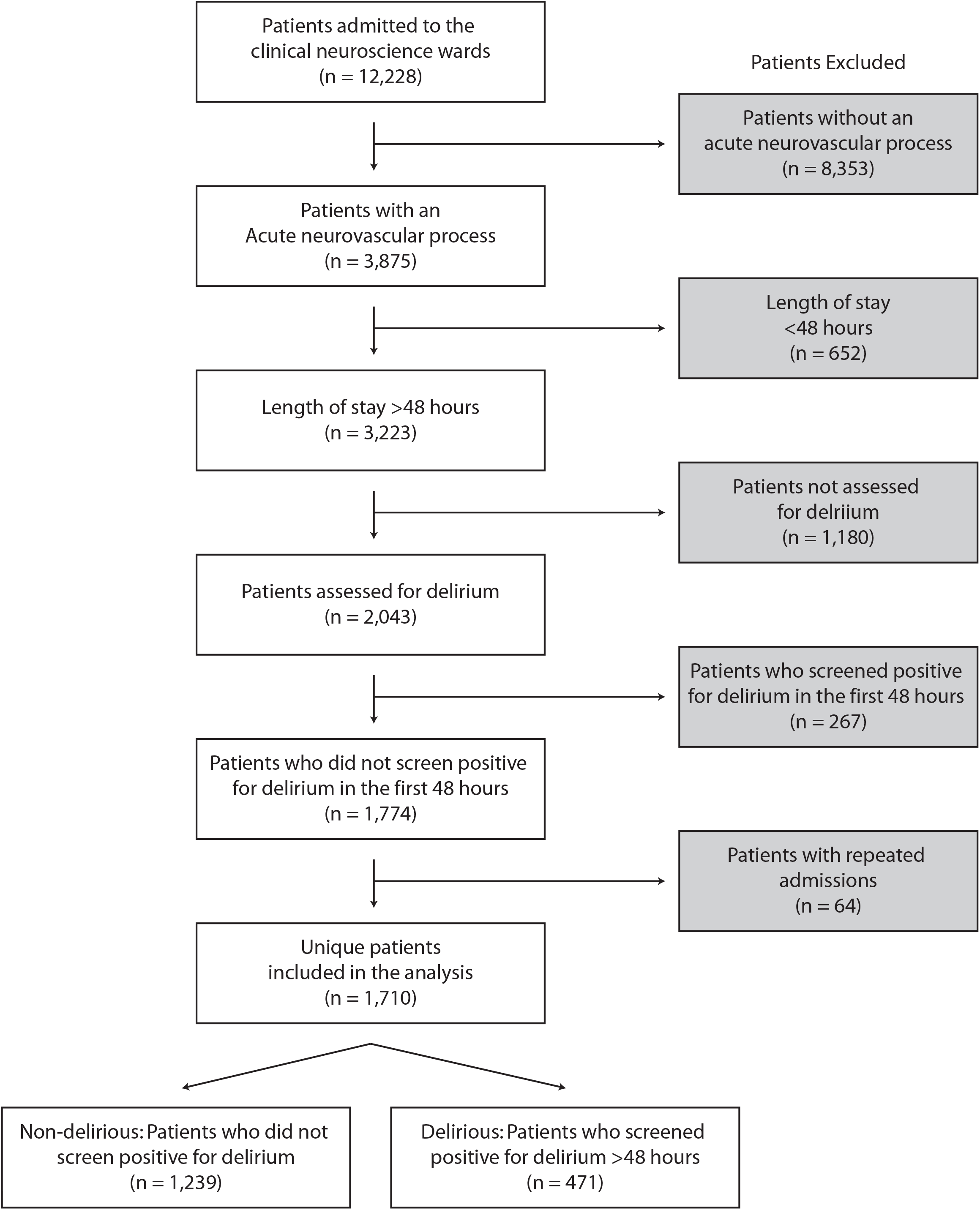
Study Design Flowchart

**Table 1.** Demographic, clinical characteristics, and health outcomes for the study cohort according to patients who did not develop delirium or who developed delirium 48 hours or more after admission. Patients assessed indicate the number of patients for whom data was potentially available for the characteristics depicted in the relevant row. Costs were determined from a hospital administrative database and can only be reported in relative units, as per hospital administrative policy.

|  | Patients who did not develop delirium |  | Patients who developed delirium |  |  |
| --- | --- | --- | --- | --- | --- |
|  | Patients assessed | # patients (%) | Patients assessed | # patients (%) | p-value* |
| **Delirium development** |  | 1239<br>(72.5% of cohort) |  | 471<br>(27.5% of cohort) | n/a |
| Time to delirium development, days, median (IQR) | 1239 | n/a | 471 | 5.65 (3.29, 10.54) | n/a |
| Num of delirious days, median (IQR) | 1239 | n/a | 471 | 3 (2, 7) | n/a |
| **Demographics** |  |  |  |  |  |
| Age, median (IQR) | 1239 | 66.95 (56.12, 77.19) | 471 | 72.28 (61.49, 81.87) | <0.0001 |
| Female sex, n (%) | 1239 | 588 (47.46%) | 471 | 203 (43.10%) | 0.11 |
| Married, n (%) | 1179 | 642 (54.45%) | 435 | 211 (48.51%) | 0.03 |
| English speaker, n (%) | 1239 | 1073 (86.60%) | 471 | 400 (84.93%) | 0.37 |
| Latinx ethnicity, n (%) | 1131 | 78 (6.90%) | 410 | 32 (7.80%) | 0.54 |
| **Race, n (%)** |  |  |  |  |  |
| White | 1238 | 932 (75.28%) | 471 | 356 (75.58%) | 0.98 |
| Black |  | 100 (8.08%) |  | 37 (7.86%) |  |
| Other |  | 206 (16.64%) |  | 78 (16.56%) |  |
| **Insurance Payer, n (%)** |  |  |  |  |  |
| Commercial | 1239 | 368 (29.70%) | 471 | 75 (15.92%) | <0.0001 |
| Medicaid |  | 157 (12.67%) |  | 72 (15.29%) |  |
| Medicare |  | 683 (55.13%) |  | 313 (66.45%) |  |
| Other |  | 31 (2.50%) |  | 11 (2.34%) |  |
| **Baseline characteristics** |  |  |  |  |  |
| Visual impairment at baseline, n (%) | 1193 | 978 (81.98%) | 436 | 372 (85.32%) | 0.11 |
| Hearing impairment at baseline, n (%) | 1137 | 575 (50.57%) | 447 | 314 (70.25%) | <0.0001 |
| Denture use at baseline, n (%) | 1239 | 126 (10.17%) | 471 | 60 (12.74%) | 0.13 |
| Memory impairment at baseline, n (%) | 403 | 108 (26.80%) | 117 | 55 (47.01%) | <0.0001 |
| No. dependent ADLs, median (IQR) | 1239 | 1.0 (0, 3) | 471 | 2.0 (0, 5) | <0.0001 |
| Ever smoker, n (%) | 1160 | 202 (17.41%) | 364 | 73 (20.05%) | 0.25 |
| Potentially hazardous alcohol use, n (%) | 218 | 71 (32.57%) | 48 | 23 (47.92%) | 0.04 |

| **Clinical characteristics** |  |  |  |  |  |
| --- | --- | --- | --- | --- | --- |
| Intracranial hemorrhage, n (%) | 1239 | 406 (32.77%) | 471 | 243 (51.59%) | <0.0001 |
| Ischemic stroke, n (%) | 1239 | 787 (63.52%) | 471 | 233 (49.47%) | <0.0001 |
| NIHSS (ischemic stroke patients only), median (IQR) | 545 | 3 (1, 8) | 164 | 12 (5, 18) | <0.0001 |
| tPA received, n (%) | 1239 | 77 (6.21%) | 471 | 49 (10.40%) | 0.003 |
| Neurovascular intervention (within first 48hrs for ischemic stroke patients), n (%) | 787 | 50 (6.35%) | 233 | 34 (14.59%) | <0.0001 |
| ICU (within first 48hrs), n (%) | 1239 | 544 (43.91%) | 471 | 383 (81.32%) | <0.0001 |
| Ventilator (within first 48hrs), n (%) | 1239 | 171 (13.80%) | 471 | 134 (28.45%) | <0.0001 |
| Restraints (within first 48hrs), n (%) | 1239 | 175 (14.12%) | 471 | 238 (50.53%) | <0.0001 |
| Urinary catheter (within first 48hrs), n (%) | 1239 | 426 (34.38%) | 471 | 323 (68.58%) | <0.0001 |
| **Clinical Outcomes** |  |  |  |  |  |
| LOS, median (IQR) | 1239 | 4.94 (3.26-8.00) | 471 | 13.66 (8.01-21.32) | <0.0001 |
| 30-day readmissions, n (%) | 1239 | 222 (17.92%) | 471 | 109 (23.14%) | 0.01 |
| Total cost, \$/constant, mean | 1236 | 2744 | 470 | 5686 | <0.0001 |
| **Discharge disposition** |  |  |  |  |  |
| Discharge to home | 1239 | 691 (55.77%) | 471 | 53 (11.25%) | <0.0001 |
| SNF/Rehab/LTC |  | 471 (38.01%) |  | 367 (77.92%) |  |
| In-hospital death |  | 41 (3.31%) |  | 26 (5.52%) |  |
| Hospice |  | 22 (1.78%) |  | 17 (3.61%) |  |
| Other |  | 14 (1.13%) |  | 8 (1.70%) |  |

\* Rank-sum test was used for continuous variables and Chi-square test or Fisher exact test were used for categorical variables.

### Acute clinical characteristics

Patients who developed delirium were more likely to have intracranial hemorrhage and less likely to have ischemic stroke than patients who did not develop delirium (Table 1). Median NIH stroke scale scores among ischemic stroke patients were higher for patients who developed delirium. Patients who developed delirium were also more likely to have received tissue plasminogen activator and neurovascular intervention, and more likely to have been in the ICU during the first 48 hours of admission and to have been on a ventilator. Patients who developed delirium were also more likely to receive physical restraints and urinary catheters within the first 48 hours of hospitalization.

### Clinical outcomes

Clinical outcomes were worse in patients who developed delirium during their hospitalization (Table 1), including longer length of stay, higher rates of 30-day readmission, higher in-hospital mortality, and increased discharge to facilities rather than home. The relative total cost of hospitalization was more than two times higher among patients who developed delirium.

### Medications classes significantly associated with delirium

Patients who developed delirium received more unique medications in the first 48 hours after admission (median 24, IQR 18-29) compared to those who never developed delirium (median 18, IQR 12-23, p < 0.0001).

### Antipsychotics

Patients who developed delirium were more likely to have received early antipsychotics (Table 2). Haloperidol was the most commonly prescribed typical antipsychotic and quetiapine the most commonly prescribed atypical antipsychotic. Both typical and atypical antipsychotics were significantly associated with delirium, even in the multivariable model (Table 3). Following delirium identification, there was a significant reduction in the use of typical antipsychotics, but a significant increase in the use of atypical antipsychotics (Table 4).

**Table 2.** Comparison of medication exposures within the first 48 hours between non-delirious and delirious patients.

|  | **Non-delirious**<br>**# patients=1239** | **Delirious**<br>**# patients=471** | **Risk Ratio [95% CI]** | **p-value\*** |
| --- | --- | --- | --- | --- |
| Antipsychotics (all) | 83 (6.70%) | 102 (21.66%) | 3.23 [2.37, 4.40] | <0.0001 |
| Typical Antipsychotics | 42 (3.39%) | 66 (14.01%) | 4.13 [2.77, 6.17] | <0.0001 |
| Atypical Antipsychotics | 52 (4.20%) | 62 (13.16%) | 3.14 [2.14, 4.60] | <0.0001 |
| Sedatives | 132 (10.65%) | 121 (25.69%) | 2.41 [1.84, 3.15] | <0.0001 |
| Cholinesterase inhibitors | 17 (1.37%) | 14 (2.97%) | 2.17 [1.06, 4.43] | 0.03 |
| Antimicrobials | 261 (21.07%) | 166 (35.24%) | 1.67 [1.34, 2.09] | <0.0001 |
| Opiates | 380 (30.67%) | 226 (47.98%) | 1.56 [1.29, 1.90] | <0.0001 |
| Antiepileptic | 439 (35.43%) | 235 (49.89%) | 1.41 [1.16, 1.70] | <0.0001 |
| Parkinson's Medications | 17 (1.37%) | 8 (1.70%) | 1.24 [0.53, 2.89] | 0.62 |
| Steroids | 117 (9.44%) | 54 (11.46%) | 1.21 [0.86, 1.70] | 0.21 |
| Benzodiazepines | 294 (23.73%) | 120 (25.48%) | 1.07 [0.85, 1.36] | 0.45 |
| Anticholinergics | 70 (5.65%) | 28 (5.94%) | 1.05 [0.67, 1.65] | 0.81 |
| Antiemetic | 319 (25.75%) | 123 (26.11%) | 1.01 [0.80, 1.28] | 0.88 |
| Sleep aids | 142 (11.46%) | 50 (10.62%) | 0.93 [0.66, 1.30] | 0.62 |
| Stimulant | 21 (1.69%) | 7 (1.49%) | 0.88 [0.37, 2.08] | 0.76 |
| Mood stabilizer | 292 (23.57%) | 96 (20.38%) | 0.86 [0.67, 1.11] | 0.16 |
| Muscle relaxants | 21 (1.69%) | 4 (0.85%) | 0.50 [0.17, 1.47] | 0.26 |

\* Chi-square test or Fisher exact test were used for categorical variables.

**Table 3.** Multivariable model testing associations of medication classes associated with the development of delirium.

|  | **Relative risk** | **95% CI** | **p-value\*** |
| --- | --- | --- | --- |
| Patient's age | 1.03 | 1.02, 1.04 | <0.0001 |
| Female | 0.73 | 0.58, 0.92 | 0.01 |
| Hemorrhage | 1.44 | 1.07, 1.93 | 0.02 |
| Ventilator used within first 48 hours | 1.06 | 0.55, 2.04 | 0.87 |
| Typical antipsychotic used within first 48 hours | 2.96 | 1.90, 4.63 | <0.0001 |
| Atypical antipsychotic used within first 48 hours | 2.29 | 1.47, 3.55 | 0.0002 |
| Sedative used within first 48 hours | 2.13 | 1.06, 4.28 | 0.03 |
| Cholinesterase inhibitor used within first 48 hours | 1.38 | 0.62, 3.09 | 0.43 |
| Antimicrobial used within first 48 hours | 1.47 | 1.12, 1.93 | 0.006 |
| Opiate used within first 48 hours | 1.68 | 1.28, 2.21 | 0.0002 |
| Antiepileptic used within first 48 hours | 0.93 | 0.69, 1.25 | 0.63 |

**Table 4.** Comparison of medications pre- and post-delirium identification for delirious patients only

|  | **Pre-delirium**<br>**(n=471)** | **Post-delirium**<br>**(n=471)** | **Absolute**<br>**Change (%)** | **p-value** |
| --- | --- | --- | --- | --- |
| **Whether the medication was taken, n (%)** |  |  |  |  |
| Antipsychotics (all) | 192 (40.76%) | 211 (44.80%) | +4.04% | 0.07 |
| Typical Antipsychotics | 101 (21.44%) | 65 (13.80%) | +7.64% | 0.0007 |
| Atypical Antipsychotics | 156 (33.12%) | 203 (43.10%) | +9.98% | <0.0001 |
| Sedatives | 140 (29.72%) | 30 (6.37%) | -23.35% | <0.0001 |
| Cholinesterase inhibitors | 17 (3.61%) | 20 (4.25%) | +0.64% | 0.18 |
| Antimicrobials | 241 (51.17%) | 208 (44.16%) | -7.01% | 0.01 |
| Opiates | 270 (57.32%) | 205 (43.52%) | -13.8% | <0.0001 |
| Antiepileptic | 259 (54.99%) | 215 (45.65%) | -9.34% | <0.0001 |
| Parkinson's Medications | 11 (2.34%) | 14 (2.97%) | +0.63 % | 0.18 |
| Steroids | 83 (17.62%) | 61 (12.95%) | -4.67% | 0.005 |
| Benzodiazepines | 176 (37.37%) | 93 (19.75%) | -17.62% | <0.0001 |
| Anticholinergics | 57 (12.10%) | 51 (10.83%) | -1.27% | 0.45 |
| Antiemetic | 183 (38.85%) | 114 (24.20%) | -14.65% | <0.0001 |
| Sleep aids | 194 (41.19%) | 263 (55.84%) | +14.65% | <0.0001 |
| Stimulant | 55 (11.68%) | 76 (16.14%) | +4.46% | 0.0004 |
| Mood stabilizer | 156 (33.12%) | 181 (38.43%) | +5.31% | 0.0007 |
| Muscle relaxants | 11 (2.34%) | 16 (3.40%) | +1.06% | 0.23 |

McNemar's test was used to compare agreement of medication used before and after delirium.

### Sedatives

Patients who developed delirium were more likely to have received early sedatives. Propofol and dexmedetomidine were the most commonly prescribed sedatives. In the multivariable model, sedatives remained significantly associated with delirium, even accounting for ventilation status. Additionally, the association between sedatives and delirium remained present even when performing a stratified analysis within just ventilated patients (Supplementary Table S3). Following delirium identification, there was a significant reduction in the use of sedatives.

### Cholinesterase Inhibitors and Memantine

Patients who developed delirium were more likely to have received early cholinesterase inhibitors or memantine. However, only 1.8% of patients received such medications in this time frame (31/1710). Donepezil and memantine were the most commonly prescribed medications in this group. However, the association was not significant in the multivariable model and there was no significant change in administration after delirium identification.

### Antimicrobials

Patients who developed delirium were more likely to have received early antimicrobials. The most commonly prescribed antimicrobials were cefazolin, vancomycin, and ciprofloxacin. In the multivariable model, antimicrobials remained significantly associated with delirium. Following delirium identification, there was a significant reduction in the use of antimicrobials.

### Opiates

Patients who developed delirium were more likely to have received early opiates. Opiates were the most commonly prescribed medication class among all patients (475/1710, 27.8%), and oxycodone and hydromorphone were the most commonly prescribed opiates. In the multivariable model, opiates remained significantly associated with delirium. Following delirium identification, there was a significant reduction in opiate administration.

### Antiepileptics

Patients who developed delirium were more likely to have received early antiepileptics. Levetiracetam was the most commonly prescribed antiepileptic medication. However, the association was not significant in the multivariable model. Following delirium identification, there was a significant reduction in antiepileptic administration.

### Medications not significantly associated with delirium

#### Anticholinergics

Oxybutynin was the most commonly prescribed anticholinergic medication. Anticholinergics were not significantly associated with the development of delirium. Following delirium identification there was no significant change in anticholinergic administration.

#### Steroids

Dexamethasone was the most commonly prescribed steroid. Steroids were not significantly associated with the development of delirium. Following delirium identification, however, there was a significant reduction in steroid administration.

#### Benzodiazepines

Lorazepam was the most commonly prescribed benzodiazepine. Benzodiazepines were not significantly associated with the development of delirium. Following delirium identification, however, there was a significant reduction in benzodiazepine administration.

#### Stimulants

Only 1.6% of patients (28/1710) received stimulants, the most commonly prescribed of which was amantadine. Stimulants were not significantly associated with the development of delirium. Following delirium identification, however, there was a significant increase in stimulant administration.

#### Sleep Aids

Sleep aids were not significantly associated with the development of delirium in the univariate analysis. Melatonin and trazodone were the most common sleep aids prescribed. Following delirium identification, however, there was a significant increase in sleep aid administration.

## DISCUSSION

Delirium is a complex, multifactorial syndrome that is common and associated with poor prognosis among patients with stroke, driving strong interest in identifying potentially modifiable risk factors to help prevent or treat it. In patients admitted to a comprehensive stroke center, several classes of medications administered in the first 48 hours were associated with the subsequent development of delirium, including antipsychotics, sedatives, opiates, and antimicrobials. Establishing a temporal context with medication administration preceding delirium is an understudied but important step to understanding the clinical significance of medications associated with delirium, as identified previously in other patient populations.

As in other studies, the association between a medication class and delirium is likely to be complex. For example, antipsychotics could be potentially deliriogenic by impairing cognition and movement.^56–58^ However, the association may also reflect confounding by indication, as anti-psychotics are often used off-label for the treatment of agitation, which can be a premonitory symptom of delirium. Prospective, randomized controlled trials have provided equivocal evidence on the utility of anti-psychotics for delirium, but meta-analyses suggest their routine use should be avoided.^59, 60^ While the use of anti-psychotics specifically for agitation is hard to track retrospectively, for the use of sedatives we were able to track ventilation status. Intriguingly, sedatives remained associated with delirium even after accounting for ventilation status, supporting the importance of interrupting sedation as early and as often as clinically appropriate.^20^

Opiates are also potentially deliriogenic,^23^ and in our data an association with delirium was maintained even after controlling for ventilator status and intracerebral hemorrhage. While some opiates, like meperidine, are thought to be highly deliriogenic, others, like hydromorphone, are perhaps less so.^23^ Prescribing practices within our institution did not yield sufficient power to detect differences within classes. The results, however, suggest that providers should prescribe opiates as a class cautiously in stroke patients at high risk for delirium. The decision is admittedly complex in that pain may also be deliriogenic,^61^ and further research is needed to balance these concerns.

Early antimicrobials were associated with the subsequent development of delirium. Antimicrobials are prescribed to treat infections, which may be deliriogenic. However, it is increasingly recognized that antimicrobials can have significant neuropsychiatric side effects, including delirium or encephalopathy.^21, 22, 62^ While antimicrobial administration may at times be critical, there is also the potential for misuse or overuse. Even for routine clinical concerns, such as aspiration^63^ or asymptomatic bacteriuria, there is heterogeneity and limited data to support the use of routine antibiotics.^64, 65^ More surprisingly, we observed that following delirium identification antimicrobials were actually deprescribed, suggesting that they were not strictly necessary. We could not determine if the specific reason for deprescribing was identification of delirium in this retrospective study. This data suggests the need for a prospective study of antibiotic practices in the setting of delirium with standardized definitions of infection, potentially enabling possible future randomized controlled trials comparing high vs. low thresholds for antibiotic treatment in this patient population.

Among pertinent negative results in our study was the lack of a clear association between anticholinergic medications and subsequent delirium. Anticholinergic medications are a class for which there seems to be plausible pathophysiologic rationale for delirium precipitation.^66^ However, the association in prior literature has been inconsistent.^31, 32, 67^ Notably, many medications with anticholinergic properties also fall into other classes, for example olanzapine is an anti-psychotic that is sometimes thought to have anticholinergic properties.^19^ We retained such medications in their most recognized therapeutic class to reflect prescribing indications.

At our comprehensive stroke center, there was significant deprescribing of potentially deliriogenic medications after delirium identification, with a smaller proportion of patients receiving benzodiazepines, steroids, opiates, anticholinergics, and sedatives after delirium identification. There was also an increase in sleep aid administration after delirium identification, likely reflecting attempts to regulate sleep-wake cycles. These findings suggest not only an awareness of delirium status on the part of the clinical staff but also efforts to reduce deliriogenic medication exposure after delirium has been identified. Whether deprescribing effectively treats delirium or whether it would be more effective with earlier implementation remains unclear and requires future study. We also observed what is likely off-label use of antipsychotics for delirium management, with a decrease in typical antipsychotics but an increase in atypical antipsychotics after delirium identification. Given that some delirium symptoms such as agitation may present safety concerns or barriers to transfer to rehabilitation facilities, delirium symptom control will likely remain an ongoing clinical concern in this patient population.

There are several limitations to our study. This study was performed at a single large, academic comprehensive stroke center. Although our center reflects the varying practices of many vascular neurologists, the results may not generalize to other settings. Approximately 1/3 of patients were not assessed for delirium, pointing to the need for ongoing efforts to improve clinical delirium assessment. However, patients not assessed tended to be younger and have shorter stays, suggesting that rates of delirium would have been lower in this group. Additionally, we specifically excluded patients who screened positive for delirium within the first 48 hours in order to clarify the temporal relationship between medications and the development of delirium, as confounding by indication is likelier when delirium presence and medication use are concurrent. More than three quarters of patients had their first positive delirium assessment recorded after 72 hours, creating at least a 24-hour buffer for most patients between early medication administration and subsequent development of delirium. While it is unknown whether the pathophysiology of hospital acquired or incident delirium differs from delirium already prevalent on admission, incident delirium as studied here is associated with longer length of stay.^68, 69^ Given the retrospective nature of the study, some covariate data was missing, for example knowledge of the baseline cognitive status of some patients, and home medications. However, most of the medications identified are not common outpatient medications. Lastly, medications were grouped by classes for sufficient statistical power. Larger studies may help identify specific medications within such classes associated with higher or lower risks of delirium.

## Conclusion

While several medications classes have been associated with concurrent delirium in various patient populations, it is unknown whether early administration of these medications are associated with subsequent delirium in patients with stroke. In a comprehensive stroke center, opiate, sedative, antimicrobial, and antipsychotic exposure were all significantly associated with the subsequent development of delirium when controlling for confounding. Additionally, there was significant deprescribing of several potentially deliriogenic medication classes in the setting of delirium identification, suggesting modifiable targets for future delirium prevention strategies in patients with stroke.

## Acknowledgements

The authors acknowledge the efforts of the Massachusetts General Hospital Neuroscience Delirium Task Force and neurology nurses.

## Supplemental Material

Supplementary Tables S1–S3

## Supplementary Material (Tables S1-S3)

**Associations between in-hospital medications and delirium in patients with stroke**

Sophia L. Ryan MD,^1, 2^ Xiu Liu MS,^1, 3^ Vanessa McKenna RN,^1^ Manohar Ghanta MS,^1^ Carlos Muniz MD MSc,^1, 4^ Rachel Renwick, PharmD,^1^ M. Brandon Westover MD PhD,^1^ Eyal Y. Kimchi MD PhD^1^

1. Department of Neurology, Massachusetts General Hospital

2. Department of Neurology, Mount Sinai Medical Center

3. Lawrence Center for Quality and Safety, Massachusetts General Hospital

4. Department of Neurology, SUNY Upstate Medical University

**Table S1.**
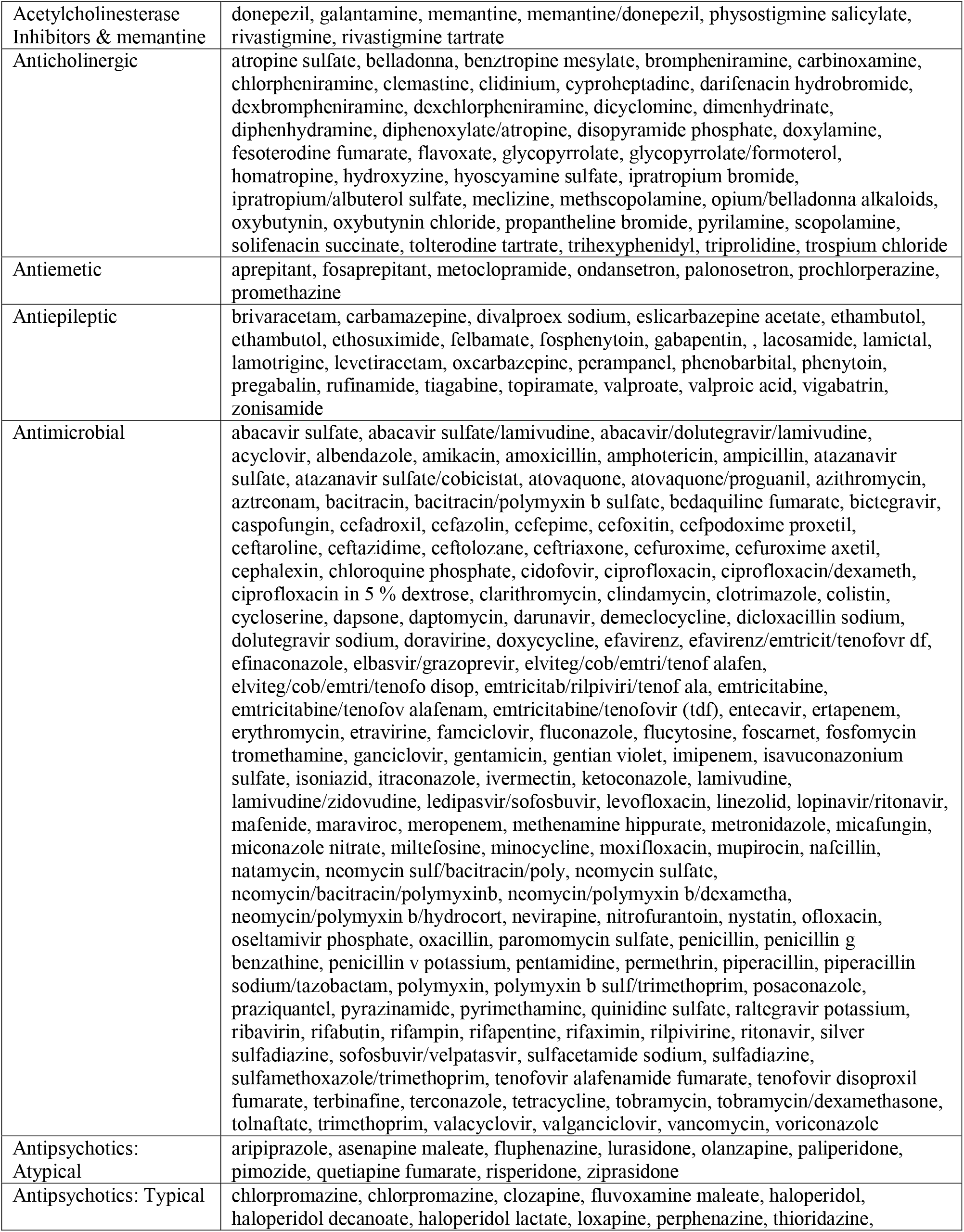
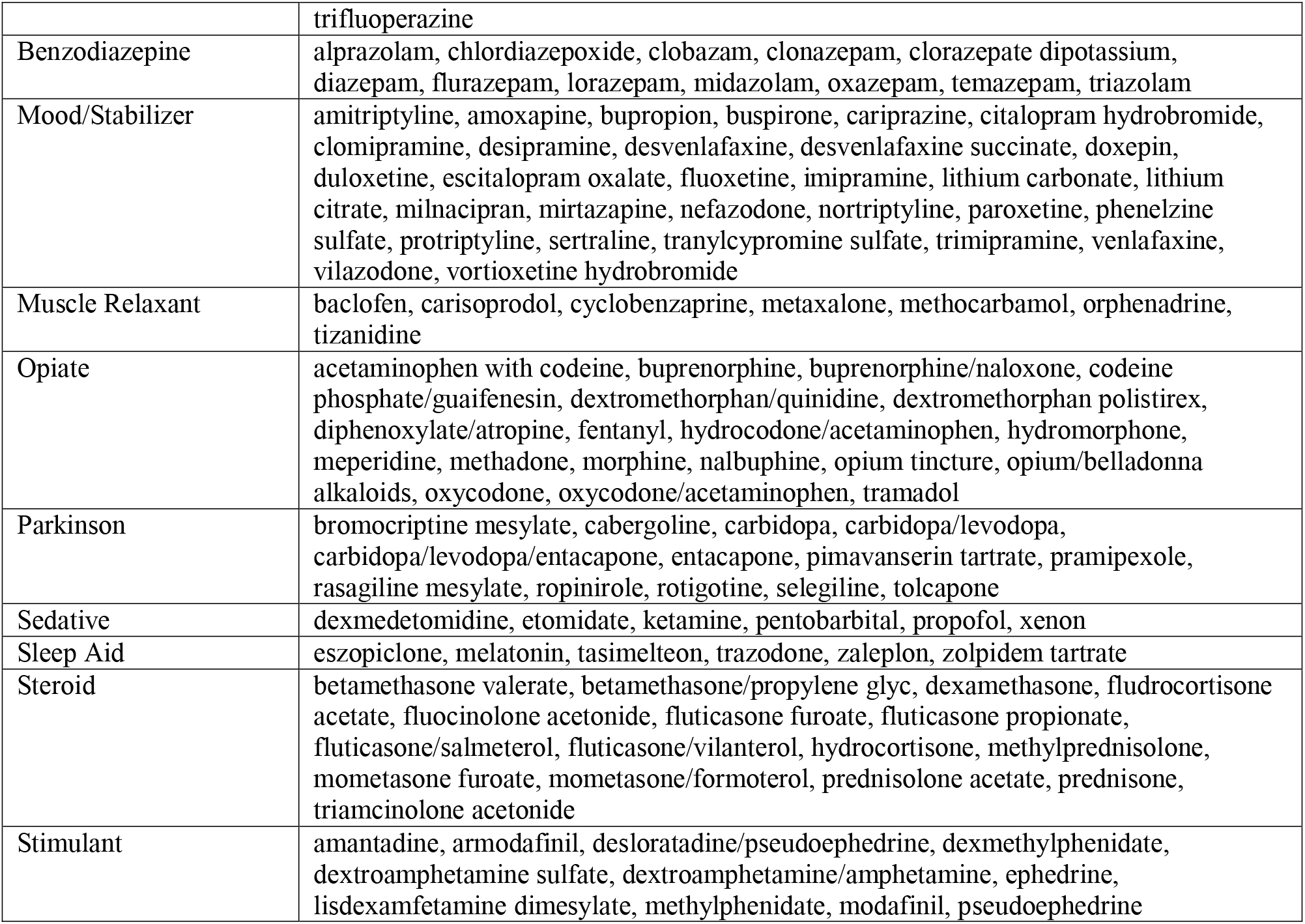
Medications included in each class

**Table S2.** Demographic, clinical characteristics, and health outcomes for the study cohort according to patients who were and were not assessed for delirium.

|  | Patients not assessed for delirium |  | Patients assessed for delirium |  |  |
| --- | --- | --- | --- | --- | --- |
|  | Patients with data | # patients (%) | Patients with data | # patients (%) | p-value* |
| **Demographics** |  |  |  |  |  |
| Age, median (IQR) | 1180 | 66.50 (55.06, 78.45) | 1710 | 68.45 (57.70, 79.03) | 0.0052 |
| Female sex, n (%) | 1180 | 545 (46.19%) | 1710 | 791 (46.26%) | 0.9700 |
| English speaker, n (%) | 1175 | 1043 (88.77%) | 1704 | 1470 (86.27%) | 0.0479 |
| Latinx ethnicity, n (%) | 1072 | 71 (6.62%) | 1551 | 110 (7.09%) | 0.6413 |
| **Race, n (%)** |  |  |  |  |  |
| White | 1107 | 915 (82.66%) | 1601 | 1296 (80.95%) | 0.2594 |
| Black | 1107 | 72 (6.50%) | 1601 | 135 (8.43%) | 0.0634 |
| Other | 1107 | 120 (10.84%) | 1601 | 170 (10.62%) | 0.8544 |
| **Baseline characteristics** |  |  |  |  |  |
| Visual impairment at baseline, n (%) | 1114 | 869 (78.01%) | 1629 | 1350 (82.87%) | 0.0015 |
| Hearing impairment at baseline, n (%) | 1046 | 486 (46.46%) | 1584 | 889 (56.12%) | <0.0001 |
| Denture use at baseline, n (%) | 1180 | 87 (7.37%) | 1710 | 186 (10.88%) | 0.0015 |
| Memory impairment at baseline, n (%) | 310 | 115 (37.10%) | 520 | 163 (31.35%) | 0.0895 |
| No. dependent ADLs, median (IQR) | 1180 | 1.00 (0.00, 3.00) | 1710 | 1.00 (0.00, 3.00) | 0.3531 |
| Ever smoker, n (%) | 1072 | 170 (15.86%) | 1524 | 275 (18.04%) | 0.1456 |
| Potentially hazardous alcohol use, n (%) | 132 | 56 (42.42%) | 266 | 94 (35.34%) | 0.1696 |
| **Clinical characteristics** |  |  |  |  |  |
| Intracranial hemorrhage, n (%) | 1180 | 472 (40.00%) | 1710 | 649 (37.95%) | 0.2670 |
| Ischemic stroke, n (%) | 1180 | 642 (54.41%) | 1710 | 1020 (59.65%) | 0.0051 |
| NIHSS (ischemic stroke patients only), median (IQR) | 425 | 3.00 (1.00, 9.00) | 765 | 4.00 (1.00, 11.00) | 0.0554 |
| tPA received, n (%) | 1180 | 61 (5.17%) | 1710 | 126 (7.37%) | 0.0182 |
| Neurovascular intervention (within first 48hrs for ischemic stroke patients), n (%) | 1180 | 32 (2.71%) | 1710 | 87 (5.09%) | 0.0016 |
| ICU (within first 48hrs), n (%) | 1180 | 530 (44.92%) | 1710 | 927 (54.21%) | <0.0001 |
| Ventilator (within first 48hrs), n (%) | 1180 | 162 (13.73%) | 1710 | 305 (17.84%) | 0.0032 |
| Restraints (within first 48hrs), n (%) | 1180 | 188 (15.93%) | 1710 | 413 (24.15%) | 0.0000 |
| Urinary catheter (within first 48hrs), n (%) | 1180 | 474 (40.17%) | 1710 | 749 (43.80%) | 0.0521 |
| **Clinical Outcomes** |  |  |  |  |  |
| LOS, median (IQR) | 1180 | 4.46 (3.04, 7.40) | 1710 | 6.26 (3.83, 12.39) | <0.0001 |
| 30-day readmissions, n (%) | 1180 | 190 (16.10%) | 1710 | 331 (19.36%) | 0.0253 |
| **Discharge disposition** |  |  |  |  |  |
| Discharge to home | 1180 | 433 (36.69%) | 1710 | 462 (27.02%) | <0.0001 |
| SNF/Rehab/LTC | 1180 | 544 (46.10%) | 1710 | 966 (56.49%) | <0.0001 |
| In-hospital death | 1180 | 87 (7.37%) | 1710 | 67 (3.92%) | <0.0001 |
| Hospice | 1180 | 41 (3.47%) | 1710 | 39 (2.28%) | 0.0545 |

\* Rank-sum test was used for continuous variables and Chi-square test or Fisher exact test were used for categorical variables.

**Table S3.**
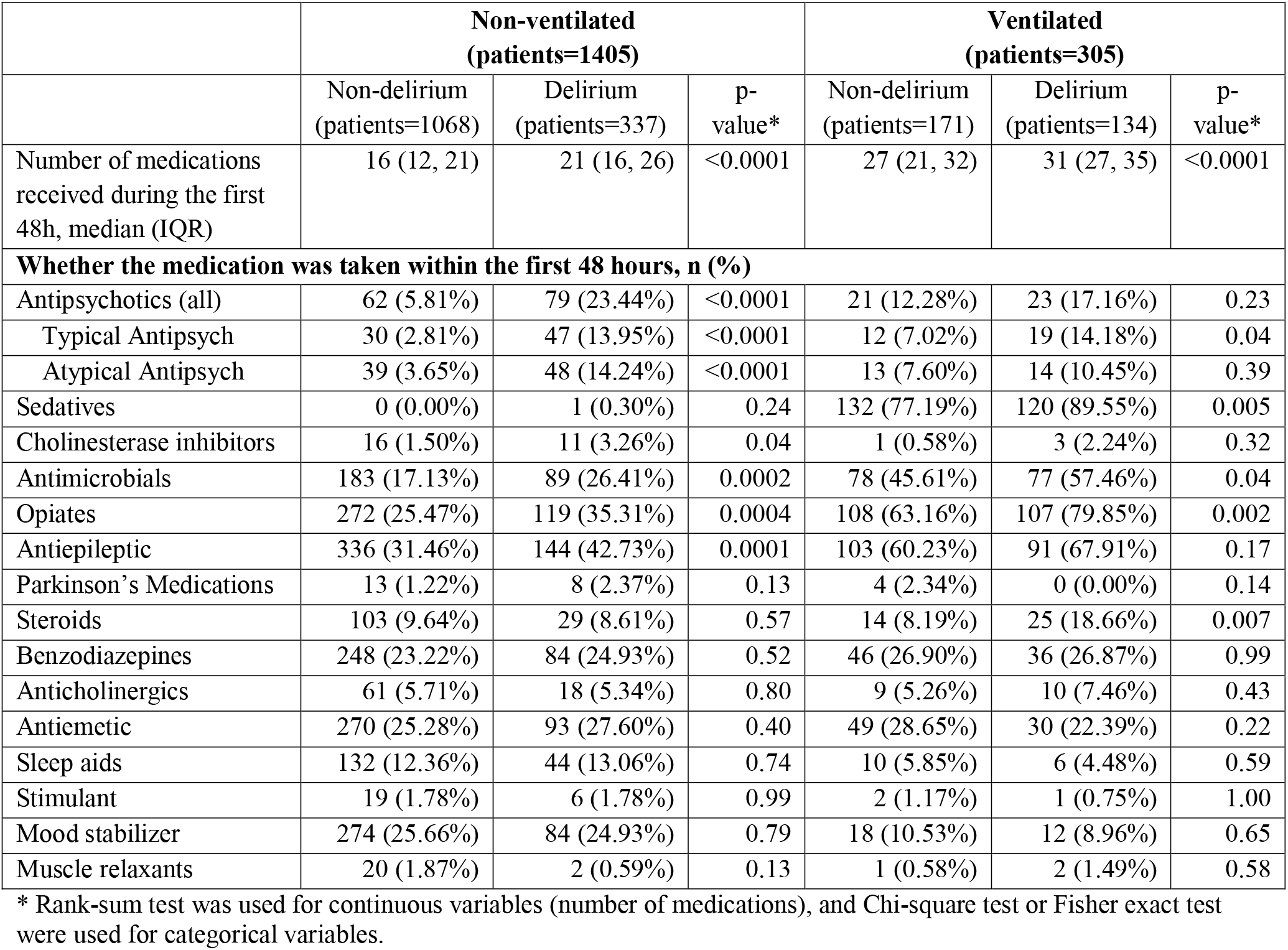
Comparison of medications used within the first 48 hours, stratified by patients’ ventilator status and whether they developed delirium. Antipsych = Antipsychotic

